# Stratifying IVF population endometria using a prognosis gradient independent of endometrial timing

**DOI:** 10.1101/2024.11.08.24316907

**Authors:** Josefa Maria Sanchez-Reyes, Antonio Parraga-Leo, Patricia Sebastian-Leon, Maria del Carmen Vidal, Diana Marti-Garcia, Katharina Spath, Immaculada Sanchez-Ribas, Francisco José Sanz, Nuria Pellicer, Jose Remohi, Dagan Wells, Antonio Pellicer, Patricia Diaz-Gimeno

**Author notes:** Josefa Maria SANCHEZ-REYES and Antonio PARRAGA-LEO are joint first authors. Corresponding author: **Patricia DIAZ-GIMENO** IVIRMA Global Research Alliance, IVI Foundation, Instituto de Investigación Sanitaria La Fe (IIS La Fe), Valencia, Spain. Edificio Biopolo – Instituto de Investigación Sanitaria la Fe Avenida Fernando Abril Martorell, 106 - Torre A, Planta 1ª, 46026 Valencia /.

## Abstract

**Background:** Independent of endometrial timing, there are molecular causes of implantation failure that disrupt the endometrial transcriptome in the mid-secretory phase. However, the molecular mechanisms disrupting the window of implantation (WOI) remain poorly understood. The molecular heterogeneity of this endometrial disruption must be characterized to develop personalized and more accurate diagnostic tools for preventive medicine, particularly for patients with a high risk of endometrial failure.

**Objective(s):** This study aimed to stratify and characterize the disrupted WOI patterns using endometrial timing-corrected whole gene expression and artificial intelligence (AI) models in *in vitro* fertilization (IVF) patients undergoing hormone replacement therapy (HRT).

**Study design:** This multicenter prospective study was conducted between January 2019 and August 2022. Endometrial biopsies were collected during the mid-secretory phase for whole endometrial transcriptome analysis using RNA-Sequencing. To identify disruptions in the WOI, the transcriptomic variation due to cyclic endometrial tissue changes was removed. Out of 195 biopsies sequenced, 131 were derived from patients that met the clinical criteria to be classified as having a poor prognosis (≥3 implantation failures, n=32) or good prognosis (<3 implantation failures, n=99). The 131 patients were randomly allocated to training (n=105) and test (n=26) sets for biomarker signature discovery and assessment of predictive performance, respectively. The reproductive outcomes of the single embryo transfer immediately after biopsy collection were analyzed. Differential gene expression and functional analyses were performed to characterize molecular profiles. Finally, a quantitative polymerase chain reaction (qPCR) assay was used to corroborate the differential expression of six potential biomarkers.

**Results:** With the dichotomous clinical classification of poor or good reproductive prognosis, there was no transcriptomic distinction between patients with a history of implantation failures during HRT endometrial preparation. Alternatively, using an AI model to stratify IVF patients based on the probability of endometrial disruption revealed molecular and clinical differences between profiles. Patients were stratified into four reproductive prognosis-related profiles, p1 (n=24), p2 (n=14), c2 (n=32) and c1 (n=61). The highest pregnancy rate (PR) was associated with c1 (91%) and the highest ongoing pregnancy rate (OPR) was associated with c2 (78%), linking these profiles to good reproductive prognoses. On the other hand, p1 had the highest biochemical miscarriage rate (BMR; 43%) while p2 had the highest clinical miscarriage rate (CMR; 43%). Notably, both p1 and p2 were related to lower PR and OPR, supporting that these profiles were associated with poor prognoses. Regarding the functional characterization in the poor prognosis profiles that were linked to miscarriages, p1 was associated with an excessive immune response against the embryo during early pregnancy stages, while p2 was initially immune-tolerant but rejected the fetus in later stages due to the lack of metabolic response.

**Conclusion(s):** This new AI-based prognostic stratification of IVF patients is promising for the clinical management of endometrial-factor infertility in precision medicine.

## INTRODUCTION

Maternal endometrial status is a key factor in successful embryo implantation and development in assisted reproductive technologies (ARTs).^1^ Cyclical physiological changes occur in the endometrium during the luteal phase to facilitate embryo implantation. Maximum uterine receptivity occurs during the window of implantation (WOI) in the mid-secretory phase.^2–5^ To ensure endometrial factor success, the endometrium must synchronize with the embryo during the WOI,^6–9^ and endometrial function must be undisrupted.^10–12^ After successful embryo implantation, the endometrium must support placentation and provide an optimal equilibrium of embryo-maternal interactions as well as adequate vascularization for fetal growth.^13^

Despite the development of ARTs, approximately 35% of transferred euploid embryos do not implant in anatomically normal uteri at the first attempt.^14^ Patients experiencing successive implantation failures are clinically classified as having recurrent implantation failure (RIF), however, there is a lack of consensus on the definition of RIF.^14,15^ The heterogeneous etiology and clinical symptoms of RIF do not provide sufficient criteria to stratify patients.^10,16^ Indeed, the molecular heterogeneity of RIF patients highlights opportunities to characterize the molecular profiles that contribute to the interpatient variability, discover new biomarkers and develop tailored treatments.

The combined use of transcriptomics and artificial intelligence (AI) algorithms has significantly advanced the understanding of endometrial-factor infertility.^10–12,17–19^ This work is laying the groundwork for new applications in precision medicine and ARTs, facilitating the characterization of reproductive diseases, patient diagnosis and prognosis. In this context, accurate patient stratification is necessary to match treatment to the right patient.^20–22^

Leveraging the use of AI algorithms, our group recently proposed a biomarker signature in luteal-phase endometrial biopsies that stratifies ART patients undergoing hormone replacement therapy (HRT) into poor or good reproductive prognosis, independent of endometrial timing.^12^ In contrast to previous studies performed in natural cycles,^10^ this prospective study included clinical follow-up to investigate if the prognostic transcriptome-based groups were related to reproductive outcomes. Despite using a 404-gene panel, dichotomous patient classification into poor or good prognosis was limited by the molecular complexity of implantation failure that requires the identification of more subtypes.^12^

Hence, the current prospective study was designed to use the whole transcriptomic profile to reproduce and refine our previous classification in a new cohort of patients undergoing HRT. Our new prognostic stratification elucidates the molecular heterogeneity of the endometrial disruptions in the mid-secretory phase, independent of endometrial timing.

## MATERIALS AND METHODS

### Ethics statement

This study was approved by the Ethics Committee of the Instituto Valenciano de Infertilidad (Valencia, Spain; 1706-FIVI-048-PD). Written informed consent was obtained from all participants.

### Participants and clinical follow-up

Participants (n=291) were recruited for a multicentric, prospective study between January 2019 and August 2022 at five private fertility clinics in Spain. Participants were scheduled for endometrial evaluation before embryo transfer due to medical indications, and met the following inclusion criteria: 18–50 years old, with a body mass index (BMI) of 19–30 kg/m^2^, undergoing HRT prior to single embryo transfer (SET) with a good-quality embryo (euploidy guaranteed by preimplantation genetic testing or oocytes from donors <35 years old), and presenting an endometrial thickness >6.5 mm with trilaminar structure in proliferative phase. Exclusion criteria were male-factor infertility (in cases with autologous sperm) as the only treatment indication, untreated reproductive pathologies that may compromise endometrial function, severe pre-menopausal symptoms, uncontrolled systemic or metabolic disorders, and co-administered medication that can interfere with ARTs.

Baseline participant characteristics were obtained from internal medical records, in accordance with data protection laws in Spain.

### Study design

IVF patients undergoing HRT were clinically classified according to their history of implantation failures. Endometrial biopsies were collected during the mid-secretory phase for RNA-Sequencing (RNA-Seq) analysis. An AI probabilistic model was developed based on the transcriptome independent of endometrial timing, avoiding biases in transcriptomic variation due to cyclical changes in the endometrium (**Supplementary Material**). The AI-determined probability of a poor prognosis was used to stratify the population. Finally, the clinical outcomes and molecular functions of the stratified profiles were studied (**Figure 1**).

**Figure 1.**
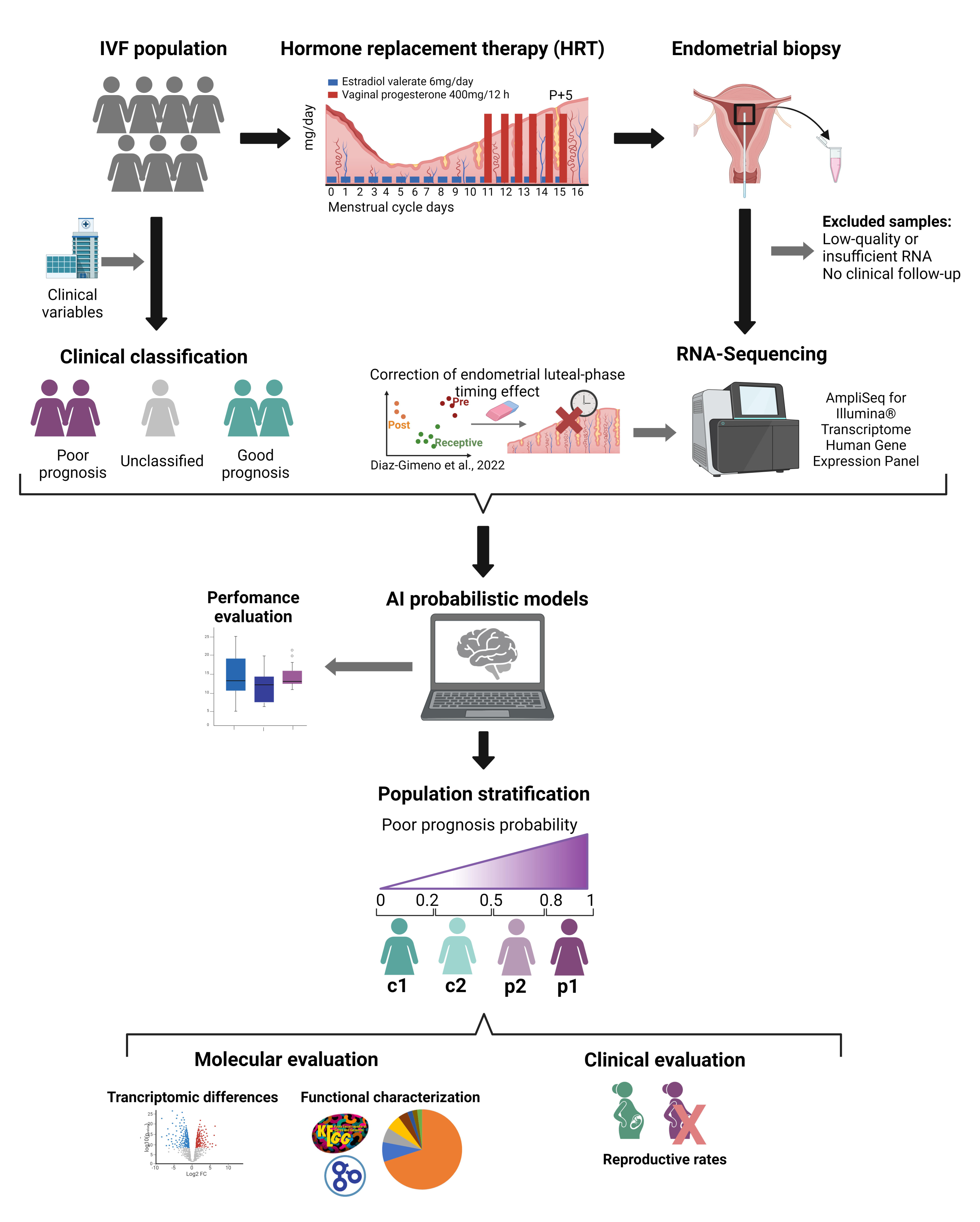
Study design. *In vitro* fertilization (IVF) patients undergoing hormone replacement therapy (HRT) were classified as having good or bad endometrial prognosis profiles based on their reproductive histories. Endometrial biopsies were processed for whole-transcriptome RNA-Sequencing. Following RNA-Sequencing data normalization, the effect of endometrial luteal-phase timing was corrected. Subsequently, an artificial intelligence model was developed to stratify the population into four groups according to their probability of having a poor prognosis. Transcriptomic evaluation and functional characterization were followed by an analysis of clinical reproductive outcomes profiles are clinically relevant.

### Endometrial biopsy collection, sequencing and data processing

Endometrial biopsies were obtained from the uterine fundus, using a cannula (Pipelle Cornier, CCD Laboratories, Paris, France) under sterile conditions, after approximately 120 hours of progesterone treatment in the HRT cycles. Anonymized samples were stored in RNA-later® (Sigma-Aldrich, Madrid, Spain) at -80°C. RNA was extracted using the miRNeasy Mini Kit following manufacturer’s instructions (Qiagen, Hilden, Germany). RNA quality was assessed using the NanoDrop™ One (AF-00342, ThermoFisher Scientific, Valencia, Spain) and 4200 TapeStation System^®^ (Agilent, Valencia, Spain). Only samples that met the following RNA quality criteria were included in the study: 260/280 ratio ∼2.0, 260/230 ratio=1.8–2.2, RNA integrity number (RIN) ≥3 and RNA fragments with more than 200 nucleotides (DV200) ≥70%.

Samples were sequenced using the AmpliSeq for Illumina^®^ Transcriptome Human Gene Expression Panel^23^ in a NextSeq 500/550 system. Raw data were evaluated using FastQC^24^. STAR^25^ was employed to align high-quality data using GRCh37/hg19 as a reference. Gene counts obtained using featureCounts^26^ were normalized using Voom transformation and quantile normalization in limma.^27^ Genes with low counts were filtered using EdgeR.^28^ Outliers and possible batch effects detected using principal component analysis (PCA) were corrected using limma linear models.^27^ Finally, the transcriptomic variation due to endometrial luteal phase timing effects was detected using a transcriptomic endometrial dating (TED) model established by our group,^19^ then removed using limma as we previously described.^12^ Additional details about the TED model are presented in **Supplementary Material**.

### Clinical classification of patients

Patients were initially classified based on their clinical history of implantation failures. Implantation failure was considered for patients who presented a negative beta chorionic gonadotropin (β-hCG) value (≤10 IU/L) 14–16 days following embryo transfer, or a biochemical miscarriage (defined by a positive serum β-hCG value, but absence of pregnancy within the first 10 weeks of gestation).^14^ Patients with at least three implantation failures were initially classified as having an endometrium with a poor prognosis whereas patients who achieved implantation success within the first three attempts, were initially classified as having an endometrium with a good prognosis. Patients with insufficient attempts were excluded from further analyses (n=62).

### Patient stratification based on AI algorithms

A training set (80% of samples) was employed to identify a biomarker signature for endometrial disruption in the mid-secretory phase, stratify patients and develop an AI-based prediction model that was externally validated in an independent testing set (20% of samples).

The training set was used for endometrial gene signature discovery, as previously described.^19^ Briefly, genes were listed in decreasing order, based on an informativity score, using CorrelationAttributeEval^29^ in Weka.^30^ To study the predictive performance of different sets of ordered genes, five-fold cross-validation processes with 100 iterations were performed independently with support vector machine (SVM),^31^ k-nearest neighbors (kNN)^32^ and random forest (RF) algorithms^33^ using RWeka.^34^ Among all the outputs, the signature with the highest accuracy and most genes was selected and used to develop a balanced probabilistic model (**Supplementary Material**). The probabilistic model was internally evaluated through cross-validation (5-fold, 10 times) in 100 different balanced models and their performance was evaluated with the test set. The range of poor prognosis probabilities obtained by running the AI model in all samples was used to stratify the study cohort into the following good (c) and poor (p) prognosis profiles: c1 (probability≤0.2), c2 (probability>0.2 & probability<0.5), p2 (probability≥0.5 & probability<0.8), or p1 (probability≥0.8).

### Molecular characterization of the different transcriptomic profiles

The differentially expressed genes (DEGs) [False Discovery Rate (FDR)<0.05] between profiles were identified using limma. Next, the functional differences between the profiles were revealed with gene set enrichment analyses (GSEA) performed using ClusterProfiler^35^. Biological functions were obtained from Kyoto Encyclopedia of Genes and Genomes (KEGG; September 2021 version)^36^ while experimental annotated terms were obtained from Gene Ontology (GO; December 2021 version).^37^

### Remeasuring the expression of selected potential biomarkers

The expression of six DEGs was evaluated in 20 samples (five samples per transcriptomic profile) with quantitative polymerase chain reaction (qPCR) using beta-actin (*ACTB*) as a housekeeping gene. RNA was reverse transcribed into cDNA using the PrimeScript Reagent Kit (Perfect Real Time, Takara, Japan) on a Thermocycler T3000 (Biometra, Ireland). The qPCR was carried out using Power-Up SYBR Green (Thermo Fisher Scientific, MA, USA) on a StepOnePlus System (Applied Biosystems, CA, USA). The specific primer sequences (Invitrogen, Thermo Fisher Scientific, MA, USA) are presented in **Supplemental Table 1**. Relative gene expression was calculated using the ΔΔCt method^38^ and *ACTB* as a housekeeping gene.

### Statistical analysis

Rates for reproductive outcomes [i.e., pregnancy (PR), cumulative pregnancy (CPR), live birth (LBR), biochemical miscarriage (BMR) and clinical miscarriage (CMR)] were calculated as described in the **Supplementary Material**. Descriptive statistics were used to ensure homogeny of the patients’ baseline clinical characteristics. Continuous variables were presented as an overall mean ± standard deviation, whereas discrete variables were presented as counts and percentages. Statistical differences between groups were compared using the Wilcoxon rank-sum test for continuous variables and the Fisher’s exact test for discrete variables. All statistical analyses were conducted in R (version 4.0.5, 2021-03-31).^39^ Graphical results were generated with ggplot2.^40^ In all cases p<0.05 was considered statistically significant.

## RESULTS

### Transcriptomic data and clinical characterization of patients

After quality control and analysis of available clinical information (see **Supplementary Material** for details), 131 samples and 14,674 genes qualified for evaluation. Patients were clinically classified as having an endometrium associated with a poor (n=32) or good (n=99) prognosis based on the number of previous implantation failures. Both groups were homogeneous in terms of main clinical variables (e.g., number of patients, age, body mass index, infertility type and duration, endometrial dating) (**Supplemental Table 2**), ensuring that there were no potential biases in endometrial-factor transcriptomic differences. As expected, the number of transfers and implantation failures were significantly different between groups (p-value=2.20e-16) due to the clinical classification criteria used for this study. However, pairwise comparison revealed there were no significant differences between the good prognosis groups (c1 and c2). Batch effects were corrected to ensure the transcriptomic differences were related to endometrial disruption independent of endometrial timing (see **Supplementary Material**).

### Four new prognostic stratification groups for endometrial function

Participants were stratified into four profiles using a 236-gene signature and a probabilistic model that predicts endometrial profiles with 77% accuracy, 67% sensitivity and 80% specificity (see **Supplementary Material** for more details). The four profiles were established as poor (p) or good (c) according to the probability of presenting an endometrium associated with a poor prognosis: p1 (n=24) and p2 (n=14), c2 (n=32) and c1 (n=61). Clinical variables were homogenous for all stratified profiles (**Table 1**).

**Table 1.** Baseline characteristics and reproductive history of the stratified groups. Endometrial profiles (c1, c2, p2, and p1) are compared according to the main clinical variables related to reproductive outcomes. Using the transcriptomic endometrial dating (TED) model, endometria in early and late secretory (ESE;LSE) classes were grouped as displaced (Ds) while early and late mid-secretory (EMSE;LMSE) classes were grouped as on time (Ot). BMI, body mass index; N/A, not available; No., number of.; P, primary; S, secondary. ***p-value < 0.001; ^a^p-value < 0.05 in p1 vs. c1; ^b^p-value < 0.05 in p1 vs. c2; ^c^p-value < 0.05 in p2 vs. c1; ^d^p-value < 0.05 in p2 vs. c2; ^e^p-value < 0.05 in p2 vs. p1.

|  | **c1** | **c2** | **p2** | **p1** | **p-value** |
| --- | --- | --- | --- | --- | --- |
| **No. patients** | 61 | 32 | 14 | 24 | N/A |
| **Age (years)** | 40.77 ± 4.41 | 39.84 ± 4.97 | 40.14 ± 4.44 | 42.5 ± 3.56 | 0.1686 |
| **BMI (kg/m<sup>2</sup>)** | 23.62 ± 3.97 | 22.53 ± 3.47 | 22.53 ± 3.34 | 22.22 ± 3.68 | 0.3324 |
| **Infertility type** | P = 48 (82.8%)<br>S = 10 (17.2%)<br>N/A = 3 | P = 23 (79.3%)<br>S = 6 (20.7%)<br>N/A = 3 | P = 12 (92.3%)<br>S = 1 (7.7%)<br>N/A = 1 | P = 18 (85.7%)<br>S = 3 (14.3%)<br>N/A = 3 | 0.8501 |
| **Infertility duration (years)** | 2.95 ± 2.86 | 3.31 ± 3.10 | 3.58 ± 1.87 | 2.8 ± 1.7 | 0.7943 |
| **Endometrial dating (TED)** | Ds = 41 (67.2%)<br>Ot = 20 (32.8%) | Ds = 18 (56.3%)<br>Ot = 14 (43.7%) | Ds = 7 (50.0%)<br>Ot = 7 (50.0%) | Ds = 17 (70.8%)<br>Ot = 7 (29.2%) | 0.4354 |
| **No. embryo transfers** | 1.75 ± 0.85 | 1.78 ± 0.91 | 2.71 ± 1.44 | 4.21 ± 1.32 | 5E-17<br>***<br>a, b, c, d, e |
| **No. implantation failures** | 0.75 ± 0.85 | 0.81 ± 1.00 | 2.00 ± 1.75 | 3.79 ± 1.14 | 4E-22<br>***<br>a, b, c, d, e |

As expected, the number of transfers and implantation failures were significantly different between the four profiles. Profiles associated with a poor prognosis showed lower PR (29.17%) and LBR (50.00%) coupled with higher CMR (42.86%) and BMR (42.68%) compared to the good prognosis profiles (**Figure 2A**). These differences were statistically significant (p-value≤0.05) for all outcomes except CMR (**Figure 2A**). Interestingly, the p1 profile was related to a higher rate of biochemical miscarriages, the p2 profile was related to clinical miscarriages and the c1 profile was associated with the best reproductive outcomes. When groups were compared pairwise significant differences were found between the p1 and c1 groups in terms of PR (p-value=0.0011), LBR (p-value=0.0478) and BMR (p-value=0.0018); between p2 and c1 groups in terms of LBR (p-value=0.0147) and CMR (p-value=0.0478), and finally, between the p1 and c2 groups in terms of PR (p-value=0.004) (**Figure 2B**). Considering all embryo transfer attempts, the cumulative PR reached 38% for p1, 76% for p2, 81% for c2, and 93% for c1 (**Figure 2C**).

**Figure 2.**
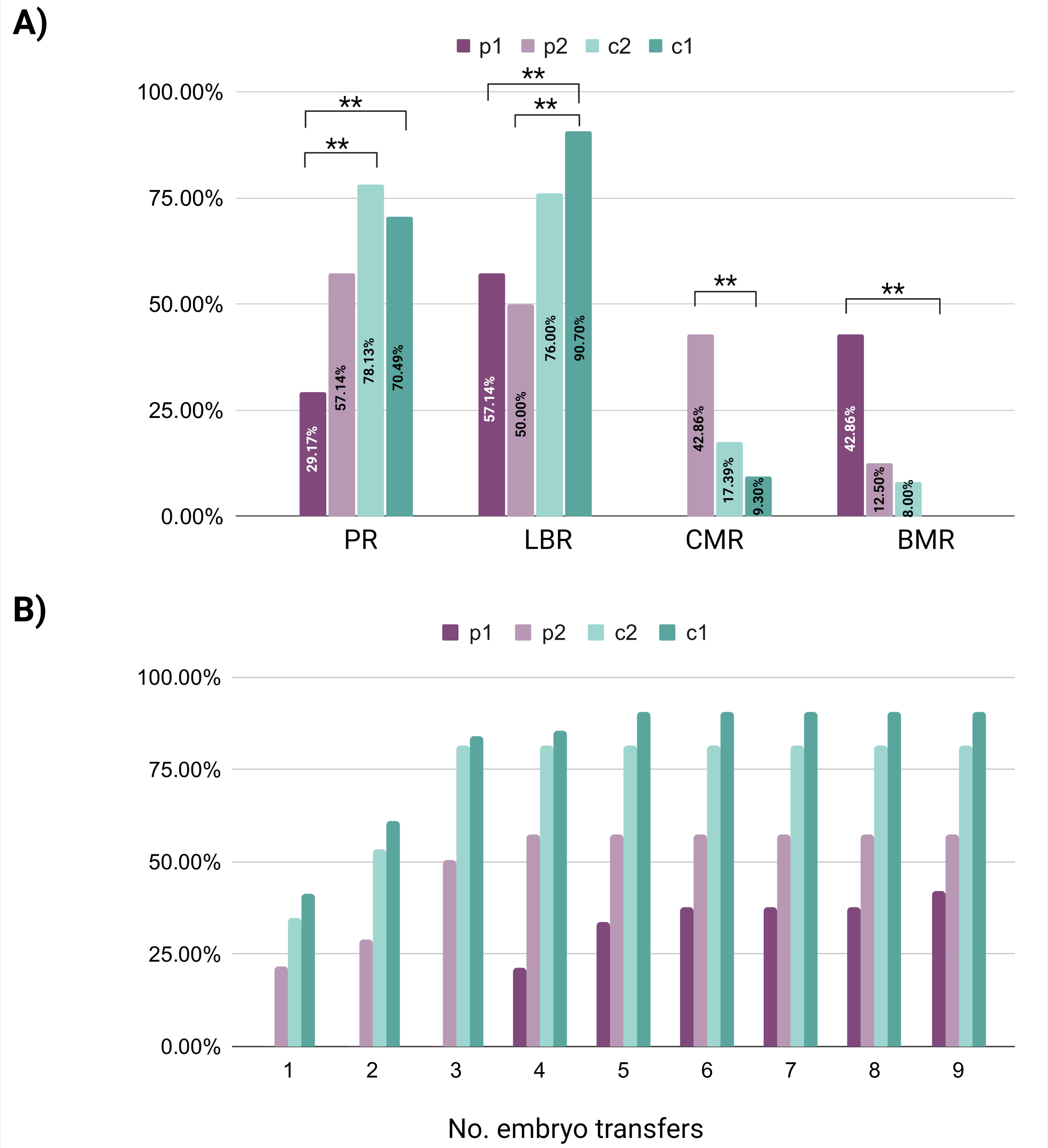
Clinical evaluation of transcriptomic profiles. **(A)** Bar plot showing the significant differences obtained through pairwise comparisons of different profiles. PR, Pregnancy rate; LBR, Live birth rate; CMR, Clinical miscarriage rate; BMR, Biochemical miscarriage rate. **(B)** Bar plot showing the cumulative pregnancy rate from multiple embryo transfers. *p-value < 0.05; **p-value < 0.01; ***p-value < 0.001.

### Molecular characterization of the transcriptomically-defined profiles

With respect to c1, there were 47 DEGs identified in p2, 3 DEGs in p1 and 1 DEG in c2. Only one transcript, mitogen-activated protein kinase 8 interacting protein 1 pseudogene (*LOC644172* or *MAPK8IP1P2*), was shared between p1 and c1 profiles, as well as c2 and c1 profiles (**Table 2**).

**Table 2.** Molecular regulation and functional differences between endometrial profiles. The table shows the number of differentially expressed genes (DEGs) and the corresponding number of significantly up-or downregulated biological functions [False Discovery Rate (FDR) < 0.05], identified through a gene set enrichment analysis (GSEA), for each comparison. The number of biological functions in each functional groups is indicated between brackets.

|  | p1 vs. c1 |  | p1 vs. c2 |  | p1 vs. p2 |  | p2 vs. c1 |  | p2 vs. c2 |  | c2 vs. c1 |  |
| --- | --- | --- | --- | --- | --- | --- | --- | --- | --- | --- | --- | --- |
| DEGs | 3 |  | 0 |  | 0 |  | 47 |  | 0 |  | 1 |  |
| Functional Groups | UP | DOWN | UP | DOWN | UP | DOWN | UP | DOWN | UP | DOWN | UP | DOWN |
| Immune response (n=26) | 9 | 0 | 9 | 0 | 7 | 1 | 3 | 10 | 6 | 2 | 0 | 9 |
| Nervous system and sensory perception (n=6) | 6 | 0 | 1 | 1 | 1 | 1 | 6 | 0 | 2 | 0 | 5 | 0 |
| Signal transduction (n=7) | 3 | 2 | 2 | 2 | 0 | 0 | 2 | 0 | 0 | 0 | 1 | 0 |
| Metabolism and energy production (n=26) | 3 | 2 | 3 | 3 | 0 | 2 | 1 | 12 | 3 | 6 | 0 | 0 |
| Cellular movement and ciliary processes (n=8) | 2 | 6 | 0 | 0 | 0 | 0 | 0 | 3 | 0 | 2 | 0 | 1 |
| Cellular adhesion and membranes (n=11) | 1 | 0 | 0 | 0 | 3 | 2 | 3 | 3 | 3 | 2 | 3 | 0 |
| Gene expression and protein degradation (n=13) | 1 | 0 | 1 | 0 | 2 | 4 | 6 | 2 | 6 | 2 | 0 | 3 |
| Proliferation and differentiation (n=2) | 1 | 1 | 1 | 0 | 0 | 0 | 0 | 0 | 0 | 0 | 0 | 0 |
| Longevity and senescence (n=2) | 1 | 0 | 0 | 0 | 0 | 0 | 1 | 0 | 0 | 0 | 0 | 0 |
| Angiogenesis and coagulation (n=3) | 0 | 0 | 0 | 0 | 0 | 2 | 2 | 0 | 1 | 0 | 0 | 0 |
| TOTAL NUMBER OF DYSREGULATED FUNCTIONS | 27 | 11 | 17 | 6 | 13 | 12 | 24 | 30 | 21 | 14 | 9 | 13 |
|  | 38 |  | 23 |  | 25 |  | 54 |  | 35 |  | 22 |  |

The p2 and p1 profiles showed the highest number of functional dysregulations compared to c1 (54 and 38 dysregulations, respectively). The downregulated functions (30/54, 55.6%) in p2 were mainly related to immune response (n=10) or metabolism and energy production (n=12). The upregulated functions (24/54, 44.4%) in p2 were mainly related with nervous system and sensory perception (n=6), gene expression and protein degradation (n=6). Compared to the c1 profile, the p1 profile had mainly upregulated functions (27/38, 71.1%), which were related to immune responses (n=9), nervous system and sensory perception (n=6). Notably, most of the downregulated functions (11/38, 28.9%) in p1 were related to cellular movement and ciliary processes (n=6) (**Table 2**).

Both poor-prognosis-related profiles had a noticeable dysregulation of immune responses compared to the c1 profiles. However, the p1 was characterized by nine upregulated functions while the p2 was characterized by ten downregulated functions. The p2 profile also presented 12 downregulated functions related to metabolism and energy production (**Table 2**).

There were 22 functional dysregulations between the control profiles, with the c2 presenting a dysregulated profile similar to p2. Specifically, the c2 profile presented a downregulation of nine functions related to immune responses and an upregulation of five functions related to the nervous system and sensory perception (**Table 2**).

### Remeasuring expression of potential biomarkers of endometrial disruption

The expression of six DEGs was validated by qPCR. Three of these DEGs were identified in the p1 vs. c1 comparison [DND microRNA-mediated repression inhibitor 1 (*DND1*), synaptotagmin 10 (*SYT10*) and mitogen-activated protein kinase 8 interacting protein 1 pseudogene (*LOC644172)*] or c2 vs. c1 comparison (*LOC644172*). The remaining three DEGs were selected for having the highest absolute fold change between p2 and c1 [CF transmembrane conductance regulator (*CFTR*), V-set domain containing T cell activation inhibitor 1 (*VTCN1*) and solute carrier family 17 member 8 (*SLC17A8*)]. Except for *SLC17A8* (**Supplemental Figure 4**), all genes showed the same gene expression trends in qPCR and RNA-Seq, reinforcing their role as potential biomarkers of endometrial disruption.

## COMMENT

### Principal findings

This study is the first to stratify endometrial function into four transcriptomic profiles, independent of endometrial timing. The four transcriptomic profiles corresponded with significantly different reproductive rates, showing a gradient of prognoses and highlighting the complex nature of endometrial disruption in the mid-secretory phase.

### Results in the context of what is known

Disrupted endometrial function, independent of luteal phase endometrial timing, was associated with a heterogeneous transcriptomic behavior among IVF patients undergoing HRT, as previously reported in patients undergoing natural cycles.^10^ Our binary prediction model (good vs. poor prognosis) was based on a signature of 236 genes that characterized the transcriptomic behavior with 67% sensitivity. Our model’s predictive performance exceeds that of Koot’s binary model, which was based on a larger signature of 301 genes and reached a 58.3% sensitivity when used to compare control and RIF patients in natural cycles.^10^ Although we improved the predictive performance, these results showed that using a dichotomous model to identify the gene expression patterns associated with good or poor prognosis did not achieve sufficient power. Our last study in patients undergoing HRT showed that stratifying patients using a 404-gene panel instead of clinical criteria significantly enhanced the transcriptomic and clinical differences between poor and good prognosis profiles.^12^ However, the probabilistic model was not tested in an independent test set, impeding comparison of the model’s performance. Thus, in this study, the whole transcriptome was used to stratify patients into four profiles according to an AI-computed probability of endometrial disruption. Notably, the resulting gradient of prognoses distinguished two profiles that were related to different types of miscarriages.

### Clinical and research implications

This work characterized four new prognostic profiles that can be used to predict reproductive outcomes in IVF patients with a history of implantation failures. Specifically, the p1 transcriptomic profile was related to the worst prognosis, characterized by the highest BMR and the lowest PR. On a molecular level, the p1 was associated with an excessive immune response, suggesting that a poor feto-maternal tolerance could be driving miscarriages in the early stages of pregnancy.^41^ Moreover, the overall downregulation of ciliary processes in this cohort of patients with RIF reinforces their role in uterine disorders.^42^ Alternatively, the p2 profile was related to the highest CMR and a lack of immune and metabolic responses. These findings suggest that implantation is facilitated by an initial immune tolerance but miscarriage occurs in subsequent pregnancy stages due to energetic deficiencies. This novel hypothesis about the relevance of the endometrial factor in clinical miscarriages requires further investigation. Finally, there were two good prognosis profiles characterized by high PRs and LBRs. The c1 profile was related to the highest LBR and lowest BMR.

Overall, this new taxonomy can help improve the precision of diagnosis and treatment of infertile women. It lays the groundwork for a new generation of tools for evaluating patients with suspected endometrial-factor infertility or stratifying types of miscarriages. Additionally, the molecular and functional differences between the reproductive prognosis profiles set the foundation for the discovery of new biomarkers and/or tailored therapeutic targets for each specific transcriptomic profile.

### Strengths and limitations

This study proposed a novel stratification based on four whole-transcriptome-based profiles with a gradient of reproductive prognosis for IVF patients undergoing HRT, improving the results from the binary classification obtained in our previous studies.^12^ Additionally, this approach leverages AI-computed probabilities which are more objective and robust than traditional approaches to classify patients with endometrial-factor infertility based on the number of implantation failures.

However, it is worth mentioning that due to the stratification into four groups and the limited sample size by group, the AI model needs further optimization. Further studies with larger patient cohorts are required to boost the statistical power of the model for population inference, assess inter-cycle reproducibility, and conduct rigorous prospective clinical trials prior to clinical implementation.^43^

### Conclusions

Regardless of endometrial timing, the heterogeneous endometrial function can be leveraged to stratify IVF patients undergoing HRT. This study uncovers four distinct prognostic groups reflecting disrupted molecular profiles related with the highest BMR and CMR (p1 and p2, respectively) or less disrupted profiles associated with the highest LBR (c1) and the highest PR (c2). These molecular findings were linked to functional differences, highlighting overactive immune responses in p1 and decreased metabolism in p2, and revealing potential mechanisms of actions underlying the biochemical and clinical miscarriages in this cohort. Taken together, our results support that good and poor endometrial prognoses have evident molecular and clinical differences. These findings advance the research in personalized diagnostic and therapeutic strategies in reproductive medicine, particularly for patients with endometrial-factor infertility.

## Paper presentation information

Some of the findings reported in this scientific article were presented at the 78^th^ American Society for Reproductive Medicine Scientific Congress & Expo (ASRM) held in Anaheim, California, USA, October 22-26, 2022.

## Data Availability

All data produced in the present study are available upon reasonable request to the authors.

## Acknowledgements

The authors thank the patients who participated in the study and the clinical staff who contributed to their recruitment, especially Elena Labarta, Juan Antonio García Velasco, Juan Giles, Ernesto Bosch, Agustín Ballesteros, Gema Castillón, Marcos Ferrando, Graciela Kohls, Francesca Gelosi, Laura Caracena, Marga Esbert, Isabel Llorens, Cristina Gaya, Mónica Toribio, and Fernando Quintana. We would like to thank Ester Castillo and Lourdes Fernandez from Illumina® Spain, for their technical support for sequencing. Finally, we acknowledge Raquel Amigo and Cristina Cardona from the Biobank and Genomics Unit of the IIS La Fe, respectively, for assisting biopsy storing and processing.

## SUPPLEMENTARY MATERIAL

**Supplemental Table 1. Specific primers employed for qPCR remeasurementof endometrial disruption biomarkers.**

*CFTR*, CF transmembrane conductance regulator; *DND1*, MicroRNA-mediated repression inhibitor 1; FW, forward; *MAPK8IP1P2, mitogen-activated protein kinase 8 interacting protein 1 pseudogene 2*, also known as *LOC644172*; qPCR, quantitative polymerase chain reaction; RV, reverse; *SLC17A8,* solute carrier family 17 member 8; *SYT10,* synaptotagmin; *VTCN1,* V-set domain containing T cell activation inhibitor 1.

**Supplemental Table 2. Homogeneity of the baseline characteristics in poor and good endometrial prognosis groups.**

With the transcriptomic endometrial dating (TED) model, early and late secretory (ESE;LSE) classes were grouped as displaced while early and late mid-secretory (EMSE;LMSE) classes were grouped as on time. BMI, body mass index; N/A, not available; No., number of. ***p-value < 0.001.

**Supplemental Table 3. Homogeneity of the baseline characteristics in training and test sets.** Baseline characteristics of training and test sets are shown. With the transcriptomic endometrial dating (TED) model, early and late secretory (ESE;LSE) classes were grouped as displaced while early and late mid-secretory (EMSE;LMSE) classes were grouped as on time. BMI, body mass index; N/A, not available; No., number of. *p-value < 0.05; ***p-value < 0.001.

**Supplemental Table 4. Comparison of AI model performance metrics.**

The table shows the performance metrics (accuracy, sensitivity, and specificity) for individual machine learning models (SVM, RF, kNN) and their combinations (SVM+kNN, SVM+RF, kNN+RF). kNN, k-Nearest neighbors; RF, Random forest; SVM, Support vector machine.

**Supplemental Figure 1. Principal component analysis (PCA) results. (A)**

PCA plot identifying two outliers (V10 and Bi16), which were excluded from subsequent analyses. **(B)** PCA plots depicting the batch effect from the sequencing run before and after correction. **(C)** PCA plots depicting the endometrial timing effect obtained using the 73-gene TED signature before and after correction. EMSE, early mid-secretory; ESE, early secretory; LMSE, late mid-secretory; LSE, late secretory; PC, principal component; TED, transcriptomic endometrial dating.

**Supplemental Figure 2. Exploratory analysis of RNA quality parameters.** Principal component analysis (PCA) plots for the **(A)** 260/230 ratio, **(B)** 260/280 ratio, **(C)** RNA fragments with more than 200 nucleotides (DV200), and **(D)** RNA integrity number (RIN). No batch effects were observed for these parameters.

**Supplemental Figure 3. Selection of the poor endometrial prognosis gene signature.** Graphs highlighting the maximum number of endometrial genes the **(A)** Support vector machine (SVM), **(B)** k-Nearest neighbors (kNN) and **(C)** Random Forest (RF) models can process with the highest accuracy. The orange dotted line represents the percentage obtained with an unbalanced proportion of good and poor prognosis classes.

**Supplemental Figure 4. qPCR validation of endometrial disruption biomarkers.** Comparison of gene expression fold change obtained with RNA-Sequencing (RNA-Seq) and quantitative polymerase chain reaction (qPCR) assays. Six differentially expressed genes (DEGS) were selected from **(A)** p1 vs. c1, **(B)** p2 vs. c1, **(C)** c2 vs. c1 comparisons.

