## Supplementary Material for "Stratifying IVF population endometria using a prognosis gradient independent of endometrial timing"

#### Material and methods

##### Transcriptomic endometrial dating (TED) model

Leveraging an artificial intelligence (AI) machine learning model along with a 73-gene signature, the TED model[1] classifies endometrial biopsies obtained during the mid-secretory phase into four distinct profiles (C1-C4). While profiles C2 and C3 correspond with the early and late mid-secretory phases (EMSE;LMSE) of the menstrual cycle, profiles C1 (early secretory; ESE) and C4 (late secretory; LSE) are indicative of a displaced mid-secretory phase.

##### Optimization of the supervised balanced probabilistic model

Due to the imbalanced nature of our cohort, reflected by a noticeably higher proportion of good prognosis samples compared to poor prognosis samples, and the immensity and complexity of the entire endometrial transcriptomic gene set, the majority class (good prognosis) had to be undersampled. This step is required to prevent potential biases that may lead to a poor performance of the AI algorithm (i.e., incorrectly identifying the poor prognosis patients or minor class).[2]

Following selection of the endometrial disruption signature in the training set, a balanced probabilistic model was developed through undersampling, which consists of selecting randomly from the largest group (good prognosis) the same number of samples from the smallest group (poor prognosis). For this purpose, the training set was randomly split into 100 balanced subgroups, each containing an equal number of poor and random good prognosis samples. Three algorithms were applied individually [i.e., Support vector machine (SVM), k-Nearest neighbors (kNN) and Random forest (RF)], or in pairs (i.e., SVM+kNN, SVM+RF and kNN+RF),[3] using the selected signature. This balanced probabilistic model was evaluated in the test set 100 times, to generate a prediction-probability of endometrial disruption for each sample. A mean prediction-probability threshold of 0.5 was then used to reclassify samples as having poor (≥ 0.5) or good (< 0.5) prognosis. To externally validate the optimal prediction model, the accuracy (proportion of true results in the population being tested), sensitivity (proportion of correctly identified positives) and specificity (proportion of correctly identified negatives) were calculated independently in the test set. The algorithm with the best overall performance considering the accuracy, specificity and sensitivity, was selected as the optimal prediction model for endometrial disruption.

**Rates for reproductive outcomes**

The pregnancy rate (PR) was evaluated as the proportion of clinically-confirmed pregnancies. The ongoing pregnancy rate (OPR) was determined as the number of ongoing pregnancies divided by the total number of clinically-confirmed pregnancies. The clinical miscarriage rate (CMR) was defined as the losses of ultrasound-detected pregnancies prior to 20–24 weeks of gestation over the total number of pregnancies in which a gestational sac was visualized. The biochemical miscarriage rate (BMR) was defined as the number of biochemical pregnancies (detected by positive serum β-hCG values but without visualization of the gestational sac within the first 10 weeks of gestation) over the total number of pregnancies. Finally, the cumulative pregnancy rate (cumulative PR), that represents the total number of embryo transfers required to achieve a successful pregnancy, was calculated considering the pregnancy outcomes of all embryo transfers.

#### Results

##### Clinical and transcriptomic selection

After excluding samples with insufficient tissue (n=15), low RNA quality (n=41) and dropouts (n=40), 195 samples were sequenced. Out of 195 selected samples, two outliers were detected and removed during the data quality control step (**Supplemental Figure 1A**). Moreover, 62 patients with insufficient embryo transfers for clinical classification were removed leaving a total of 131 samples eligible for stratification.

##### Batch effect corrections

To ensure the transcriptomic differences were related to endometrial disruption, all significant batch effects were corrected. In particular, batch effects due to the sequencing run (**Supplementary Figure 1B**) and endometrial timing were detected and corrected (**Supplementary Figure 1C**). No other batch effects owing to experimental variables were detected (**Supplementary Figure 2**).

##### Gene signature associated with poor prognosis endometrium

The optimal gene signature was selected after testing the performance of three AI algorithms (SVM, kNN, and RF). While SVM and RF algorithms worked best with gene signatures composed of 76 and 15 genes, respectively, the kNN algorithm resulted in a larger gene signature of 236 genes. Notwithstanding, the accuracy of predictions was 88.1% for SVM, 82.7% for RF and 79.2% for kNN (**Supplemental Figure 3**).

##### Selection of the best AI probabilistic model

Data was randomly split into training (n=105) and test (n=26) sets, which were homogeneous in terms of baseline characteristics (**Supplemental Table 3**). The optimal balanced probabilistic model was selected after analyzing the performances of each algorithm (kNN, SVM, RF) and their combinations (SVM+kNN, SVM+RF, kNN+RF). While the majority of AI models showed suitable accuracies and specificities (65–81% or 70–95% respectively), the SVM and kNN combination was more sensitive (67%), and the only model to exceed 50% accuracy (**Supplemental Table 4**).

### Supplementary Tables

**Supplemental Table 1.** Specific primers employed for qPCR evaluation of endometrial disruption biomarkers.

| Gene | Primer sequence |
|---|---|
| ***DND1*** | FW: CTCAAATTCAGCTCGCACCG<br>RV: AGAGGTGTGACTGCCCTTCC |
| ***SYT10*** | FW: AAGGCTCTGCACATCGTCA<br>RV: AACCAACAAGGCCAGTCCAC |
| ***MAPK8IP1P2*** | FW: TACGAGGCCTACAACATGCG<br>RV: GACGACATCGCCCTTGTGAT |
| ***CFTR*** | FW: GCTCCTACACCCAGCCATTT<br>RV: AGAACACGGCTTGACAGCTT |
| ***VTCN1*** | FW: GGCAAGGGGAATGCTAACCT<br>RV: GAGAAGTTGGCTCCCTGGTC |
| ***SLC17A8*** | FW: GGCATGGAGGCAACCTTACT<br>RV: CCACTCCGTTTGAGATCCCC |
| ***ACTB*** | FW: CGTACCACTGGCATCGTGAT<br>RV: GTGTTGGCGTACAGGTCTT |

*CFTR*, CF transmembrane conductance regulator; *DND1*, MicroRNA-mediated repression inhibitor 1; FW, forward; *MAPK8IP1P2, mitogen-activated protein kinase 8 interacting protein 1 pseudogene 2*, also known as *LOC644172*; qPCR, quantitative polymerase chain reaction*;* RV, reverse; *SLC17A8,* solute carrier family 17 member 8; *SYT10,* synaptotagmin; *VTCN1,* V-set domain containing T cell activation inhibitor 1; *ACTB,* beta actin.

**Supplemental Table 2.** Homogeneity of the baseline characteristics in poor and good endometrial prognosis groups.

| Clinical variables | Poor prognosis | Good prognosis | p-value |
|---|---|---|---|
| **No. patients** | 32 | 99 | N/A |
| **Age (years)** | 41.94 ± 4.20 | 40.42 ± 4.50 | 0.09 |
| **BMI (kg/m$^2$)** | 22.30 ± 3.84 | 23.20 ± 3.71 | 0.19 |
| **Infertility type** | Primary = 24 (82.7%)<br>Secondary = 5 (17.3%)<br>N/A = 3 | Primary = 77 (83.7%)<br>Secondary = 15 (16.3%)<br>N/A = 7 | 1.00 |
| **Infertility duration (years)** | 2.92 ± 1.68 | 3.12 ± 2.88 | 0.62 |
| **Endometrial dating (TED)** | Displaced = 23 (74.2%)<br>On time = 9 (25.8%) | Displaced = 60 (60.6%)<br>On time = 39 (39.4%) | 0.30 |
| **No. embryo transfers** | 4.16 ± 1.19 | 1.72 ± 0.81 | 2E-16<br>*** |
| **No. implantation failures** | 3.75 ± 1.08 | 0.72 ± 0.81 | 2E-16<br>*** |

With the transcriptomic endometrial dating (TED) model, early and late secretory (ESE;LSE) classes were grouped as displaced while early and late mid-secretory (EMSE;LMSE) classes were grouped as on time. BMI, body mass index; N/A, not available; No., number of. ***p-value < 0.001.

**Supplemental Table 3.** Homogeneity of the baseline characteristics in training and test sets.

| Clinical variables | Training set | Test set | p-value |
|---|---|---|---|
| **No. patients** | 105 | 26 | N/A |
| **Clinical endometrial prognosis** | Poor = 26 (24.8%)<br>Good = 79 (75.2%) | Poor = 6 (23.1%)<br>Good = 20 (76.9%) | 1.00 |
| **Age (years)** | 40.37 ± 4.37 | 42.50 ± 4.48 | 0.04 * |
| **BMI (kg/m$^2$)** | 22.95 ± 2.84 | 23.11 ± 3.41 | 0.75 |
| **Infertility type** | Primary = 83 (79%)<br>Secondary = 14 (13.4%)<br>N/A = 8 (7.6%) | Primary = 18 (69.2%)<br>Secondary = 6 (23.1%)<br>N/A = 2 (7.7%) | 0.23 |
| **Infertility duration (years)** | 3.13 ± 2.49 | 2.86 ± 3.28 | 0.38 |
| **Endometrial dating (TED)** | Displaced = 65 (61.9%)<br>On time = 40 (38.1%) | Displaced = 18 (69.2%)<br>On time = 8 (30.8%) | 0.65 |
| **No. embryo transfers** | 2.29 ± 1.46 | 2.42 ± 1.10 | 0.31 |
| **No. implantation failures** | 1.44 ± 1.65 | 1.54 ± 1.27 | 0.40 |

Baseline characteristics of training and test sets are shown. With the transcriptomic endometrial dating (TED) model, early and late secretory (ESE;LSE) classes were grouped as displaced while early and late mid-secretory (EMSE;LMSE) classes were grouped as on time. BMI, body mass index; N/A, not available; No., number of. *p-value < 0.05; ***p-value < 0.001.

**Supplemental Table 4.** Comparison of AI model performance metrics.

| Parameters | Algorithms | | | | | |
|---|---|---|---|---|---|---|
| | SVM | kNN | RF | SVM+kNN | SVM+RF | kNN+RF |
| Accuracy (%) | 81 | 69 | 65 | 77 | 81 | 69 |
| Sensitivity (%) | 33 | 50 | 50 | 67 | 33 | 50 |
| Specificity (%) | 95 | 75 | 70 | 80 | 95 | 75 |

The table shows the performance metrics (accuracy, sensitivity, and specificity) for individual machine learning models (SVM, RF, and kNN) and their combinations (SVM+kNN, SVM+RF, kNN+RF). kNN, k-Nearest neighbors; RF, Random forest; SVM, Support vector machine.

### Supplementary Figures

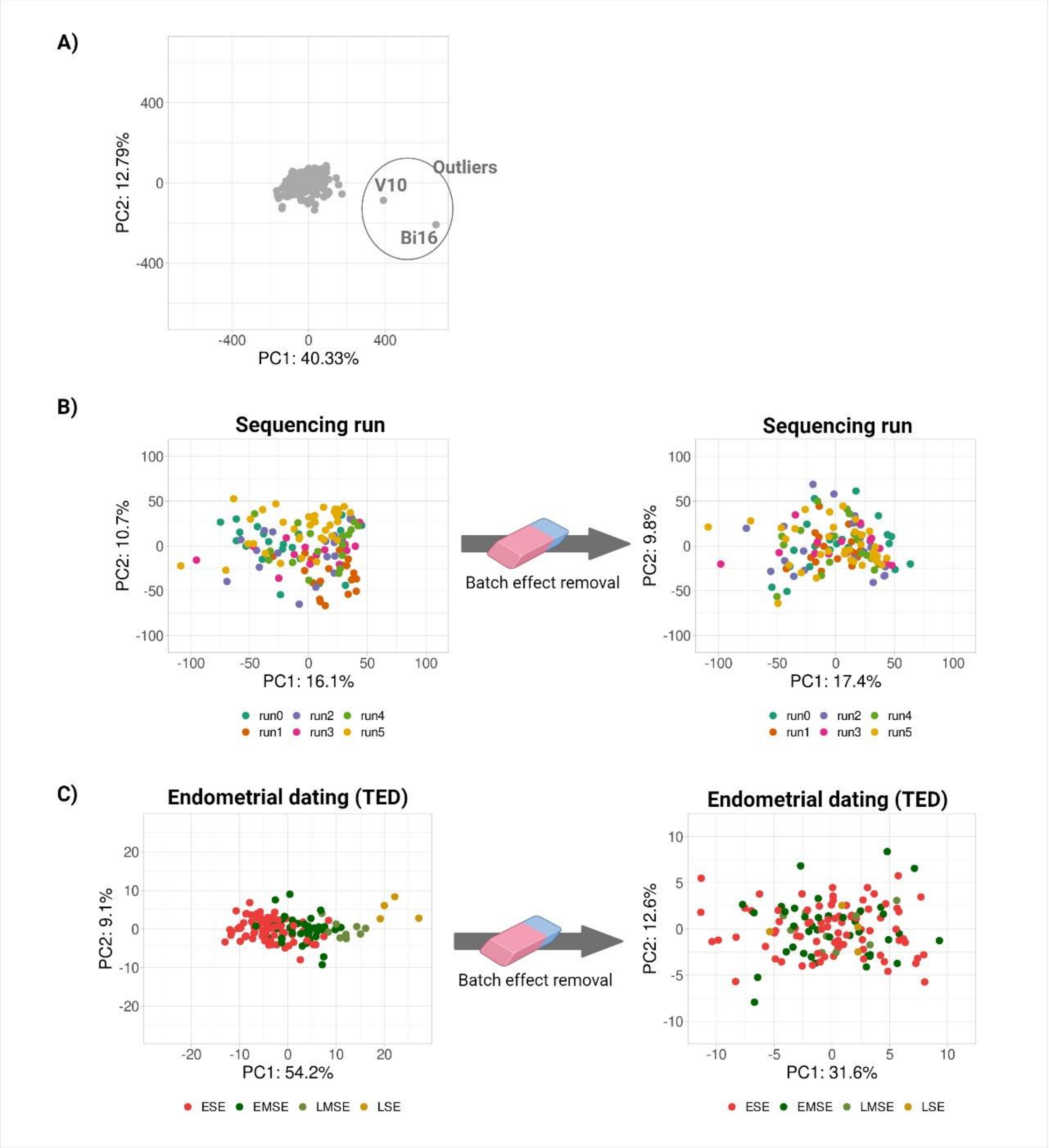


**Supplemental Figure 1. Principal component analysis (PCA) results. (A)** PCA plot identifying two outliers (V10 and Bi16), which were excluded from subsequent analyses. **(B)** PCA plots depicting the batch effect from the sequencing run before and after correction. **(C)** PCA plots depicting the endometrial timing effect obtained using the 73-gene TED signature before and after correction. EMSE, early mid-secretory; ESE, early secretory; LMSE, late mid-secretory; LSE, late secretory; PC, principal component; TED, transcriptomic endometrial dating.

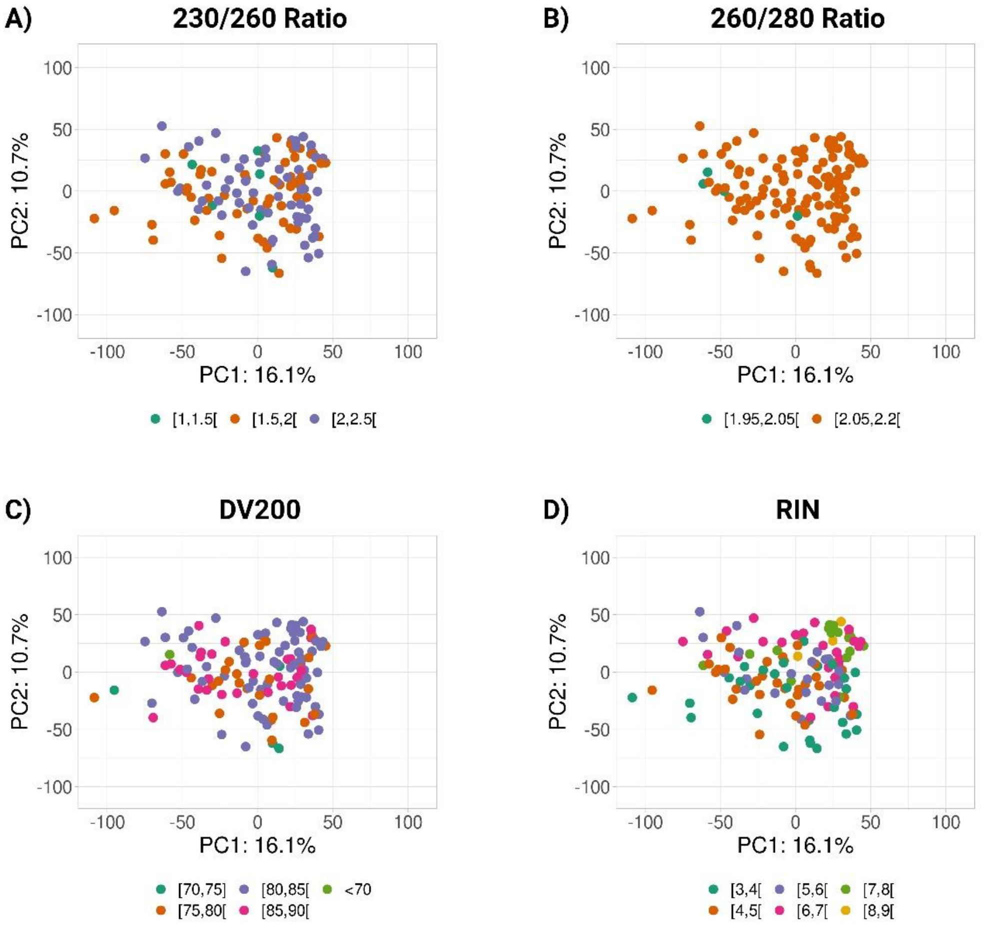


**Supplemental Figure 2. Exploratory analysis of RNA quality parameters.** Principal component analysis (PCA) plots for the **(A)** 260/230 ratio, **(B)** 260/280 ratio, **(C)** RNA fragments with more than 200 nucleotides (DV200), and **(D)** RNA integrity number (RIN). No batch effects were observed for these parameters.

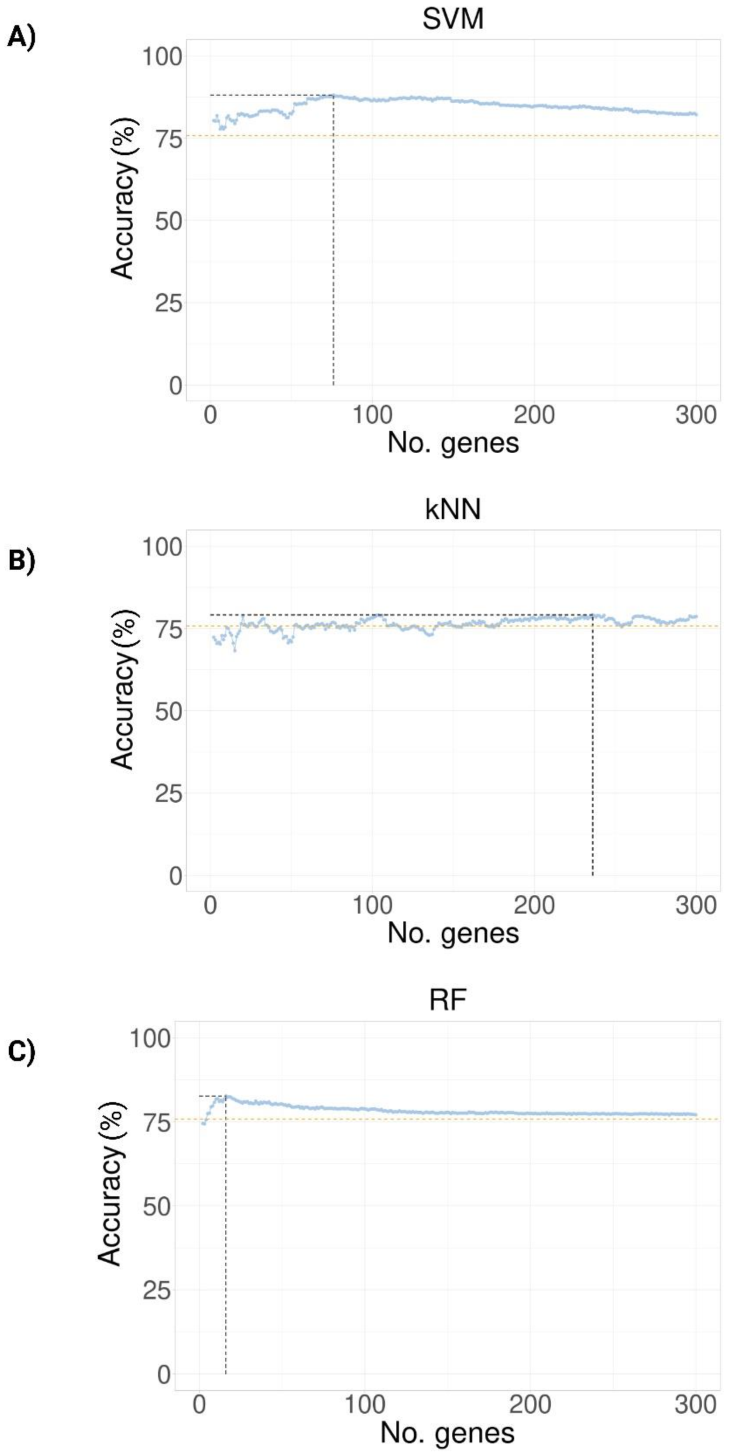


**Supplemental Figure 3. Selection of the poor endometrial prognosis gene signature.** Graphs highlighting the maximum number of endometrial genes the **(A)** Support vector machine (SVM), **(B)** k-Nearest neighbors (kNN) and **(C)**

Random Forest (RF) models can process with the highest accuracy. The orange dotted line represents the percentage obtained with an unbalanced proportion of good and poor prognosis classes.

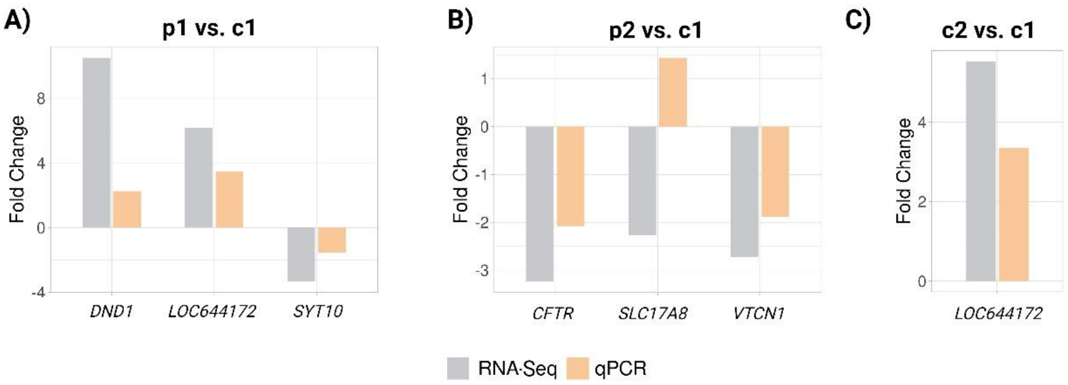


**Supplemental Figure 4. qPCR validation of endometrial disruption biomarkers.** Comparison of gene expression fold change obtained with RNA-Sequencing (RNA-Seq) and quantitative polymerase chain reaction (qPCR) assays. Six differentially expressed genes (DEGS) were selected from **(A)** p1 vs. c1, **(B)** p2 vs. c1, **(C)** c2 vs. c1 comparisons.
